# ACE2 Expression in Kidney and Testis May Cause Kidney and Testis Damage After 2019-nCoV Infection

**DOI:** 10.1101/2020.02.12.20022418

**Authors:** Caibin Fan, Kai Li, Yanhong Ding, Wei Lu, Jianqing Wang

**Affiliations:** Department of Urology, The Affiliated Suzhou Hospital of Nanjing Medical University; School of Nursing, Suzhou Vocational Health and Technical College

**Keywords:** 2019-nCoV, kidney, testis, ACE2

## Abstract

In December 2019 and January 2020, novel coronavirus (2019-nCoV) - infected pneumonia (NCIP) occurred in Wuhan, and has already posed a serious threat to public health. ACE2 (Angiotensin Converting Enzyme 2) has been shown to be one of the major receptors that mediate the entry of 2019-nCoV into human cells, which also happens in severe acute respiratory syndrome coronavirus (SARS). Several researches have indicated that some patients have abnormal renal function or even kidney damage in addition to injury in respiratory system, and the related mechanism is unknown. This arouses our interest in whether coronavirus infection will affect the urinary and male reproductive systems. Here in this study, we used the online datasets to analyze ACE2 expression in different human organs. The results indicate that ACE2 highly expresses in renal tubular cells, Leydig cells and cells in seminiferous ducts in testis. Therefore, virus might directly bind to such ACE2 positive cells and damage the kidney and testicular tissue of patients. Our results indicate that renal function evaluation and special care should be performed in 2019-nCoV patients during clinical work, because of the kidney damage caused by virus and antiviral drugs with certain renal toxicity. In addition, due to the potential pathogenicity of the virus to testicular tissues, clinicians should pay attention to the risk of testicular lesions in patients during hospitalization and later clinical follow-up, especially the assessment and appropriate intervention in young patients’ fertility.

## Introduction

Since December 2019, a novel coronavirus-induced pneumonia was discovered in Wuhan, Hubei Province, China [1, 2]. The virus is a previously unknown sub-coronal virus (β-round virus) named 2019-nCoV by WHO, which forms a branch in the subgenus sarbecvirus, subfamily Orthocoronavirinae. 2019-nCoV is closely related to SARS-CoV with above 85% identity [3]. In addition to common respiratory symptoms such as cough and fever, some patients may also experience other symptoms such as diarrhea and liver damage [4], which brings more challenges to the patient’s recovery. Although the source of the 2019-nCoV is still unknown, previous research has shown that the receptor binding domain of 2019-nCoV was able to bind ACE2 protein on the surface of human cells, which provide strong evidence for human ACE2 being the receptor for 2019-nCoV [5, 6].

Angiotensin converting enzyme 2 (ACE2) belongs to the angiotensin-converting enzyme family of dipeptidyl carboxydipeptidases, which is homologous to human angiotensin 1 converting enzyme. The expression distribution of ACE2 suggests that it might play critical roles in the regulation of cardiovascular and renal function, as well as fertility. Since the global outbreak of SARS in 2003, numerous studies have revealed the role of cell surface ACE2 as the cellular receptor for SARS-Cov and NL63 [7-9]. ACE2 has also been proven to be a major receptor of the novel 2019-nCoV because 2019-nCoV is closely related to SARS-CoV. As the virus enters the cell by binding to cell receptors to complete intracellular replication, virus release, and induce cytotoxicity, the route of virus infection depends on the expression and distribution of the corresponding receptor [10-12]. Meanwhile, the damage caused by the virus in different organs is closely related to clinical manifestations and has a major implication for understanding the pathogenesis and designing therapeutic strategies in clinical practice.

Here in this study, we analyzed the online datasets to uncover the expression pattern of ACE2 in urinary and male reproductive systems, which is the potential mechanism of abnormal renal function or even kidney damage in patients infected with 2019-nCoV. Moreover, we emphasized high ACE2 expression level in testis because of the potential pathogenicity of the virus to testicular tissues, especially the potential risks affecting fertility.

## Materials and Methods

### Clinical data

We summarized the clinical data of three previous studies to extract the incidence of abnormal renal function or kidney damage in patients infected with 2019-nCoV [2, 13, 14]. All clinical data are directly obtained from the articles above.

### Publicly available scRNA-seq gene expression data sets

In this article, we made use of some online renal single cell RNA-seq (scRNA-seq) gene expression data sets that were publicly usable. These contained the data reported in GSE131685 and GSE107585.

We used RNA and protein expression data of ACE2 in different human tissues and cancer cell lines through The Human Protein Atlas portal (Website: http://www.proteinatlas.org/) [15], GTEx portal (Website: https://gtexportal.org) and The Cancer Cell Line Encyclopedia (CCLE) [16]. All data are available directly online.

### Analysis of scRNA-seq raw sequencing data

#### Primary analysis of scRNA-seq raw sequencing data

Raw reads were processed to generate gene expression matrices as described previously. Reads with the same cell barcode, UMI and gene were grouped together to generate the number of UMIs per gene per cell. Cell number was then determined based on the inflection point of the number of UMI versus sorted cell barcode curve.

## Results

To determine whether patients infected with 2019-nCoV have abnormal renal function or kidney damage, we reviewed the latest 3 studies focused on the clinical features of such patients. In these 3 cohorts, one was a familial cluster of six patients, while other cohorts contained 99 patients and 41 patients, respectively. Outcome in two relatively larger sample studies suggest that about 3% to 10% of patients infected with 2019-nCoV had abnormal renal function, including elevated creatinine or urea nitrogen. In addition, 7% of patients experienced acute renal impairment (Table 1). Considering the large number of infected patients, it is necessary to explore the mechanisms of abnormal renal function and to promote to take special care on such patients.

**Table 1.** Summary of the renal function characteristics of patients infected with 2019-nCoV in 3 cohorts

| Characteristics | Cohort 1 | Cohort 2 | Cohort 3 |
| --- | --- | --- | --- |
| **Patients** | n=99 | n=41 | n=6 |
| **Age, years** | 55.5 (21-82) | 49 (41–58) | 10-66 |
| **Sex** |  |  |  |
| Female | 32 (32%) | 11 (27%) | 3 |
| Male | 67 (68%) | 30 (73%) | 3 |
| **Renal Function** |  |  |  |
| **Blood urea nitrogen** |  |  |  |
| Increased | 6 (6%) | N/A | 0 |
| **Serum creatinine** |  |  |  |
| Increased | 3 (3%) | 4/41 (10%) | 2 |
| **Other renal related affairs** |  |  |  |
| Acute kidney injury | N/A | 3 (7%) | N/A |

As the virus frequently enters the cell by binding to cell receptors, and ACE2 has been proven to be one of the major receptors of 2019-nCoV in human body, we explored the online datasets to find out the expression level of ACE2 in urinary system. As we expected, data from CCLE and GTEx portal indicated that ACE2 mRNA expression level is relatively higher in kidney cells (Figure 1).

**Fig. 1.**
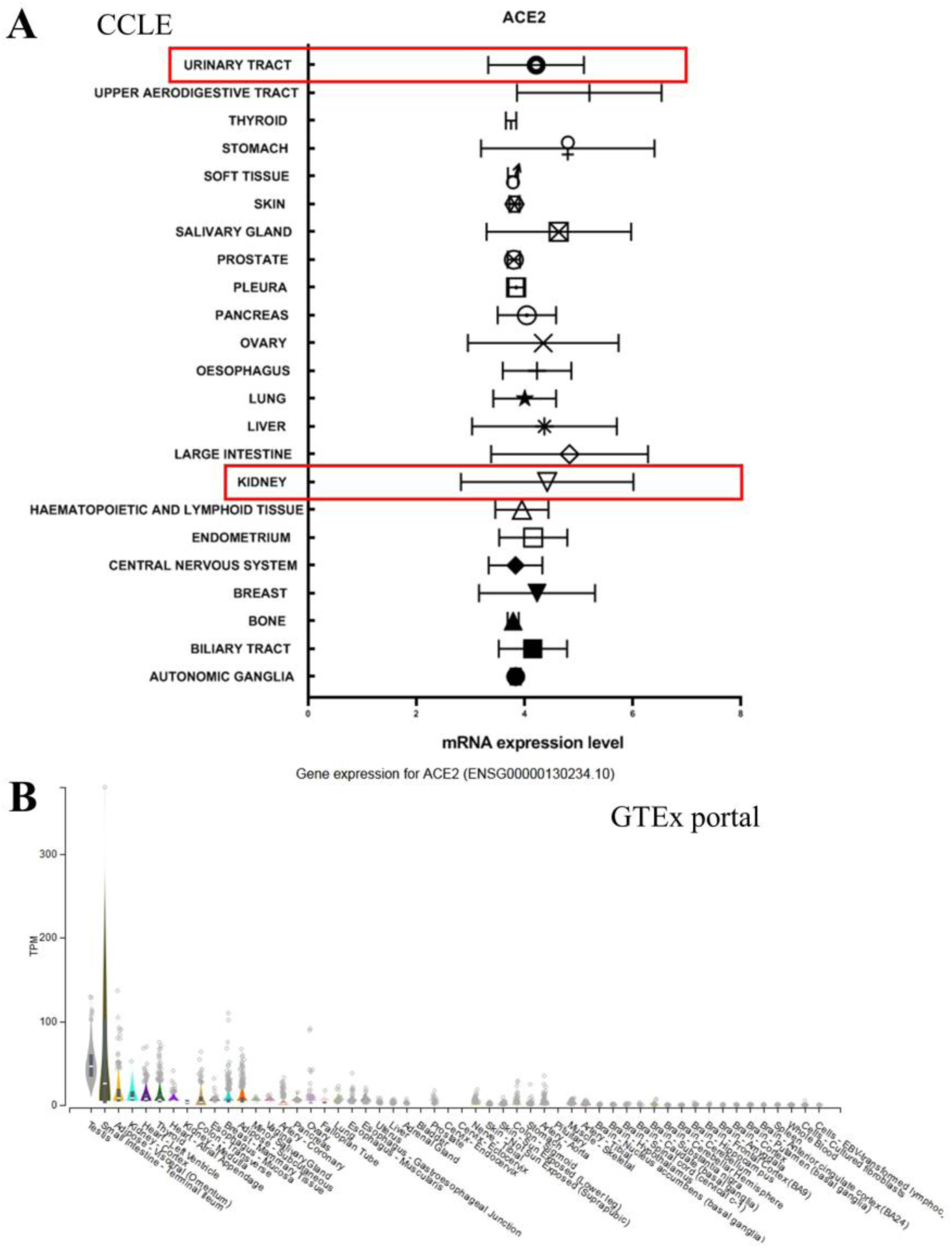
Data of mRNA expression level of ACE2 in different human tissues from online datasets. A. Data from CCLE showed ACE2 expression level in different tissues, including urinary system (red frame). B. GTEx portal showed ACE2 expression level in different tissues.

To further determine the protein expression level of ACE2 in kidney cells, we investigated the Human Protein Atlas portal to find out some details. Results of immunohistochemistry (IHC) also indicated that the expression level of ACE2 protein is significantly higher in the kidney, especially in renal tubular cells, although the mRNA expression level is not such high (Figure 2, Table 2).

Unexpectedly, we also found that ACE2 expresses quite highly in testicular cells. The protein and mRNA expression of ACE2 in the testes is almost the highest in the body. Moreover, both cells in seminiferous ducts and Leydig cells showed high ACE2 expression level (Figure 2, Table 2). These results indicate that testicular cells are the potential targets of 2019-nCoV.

**Table 2.**
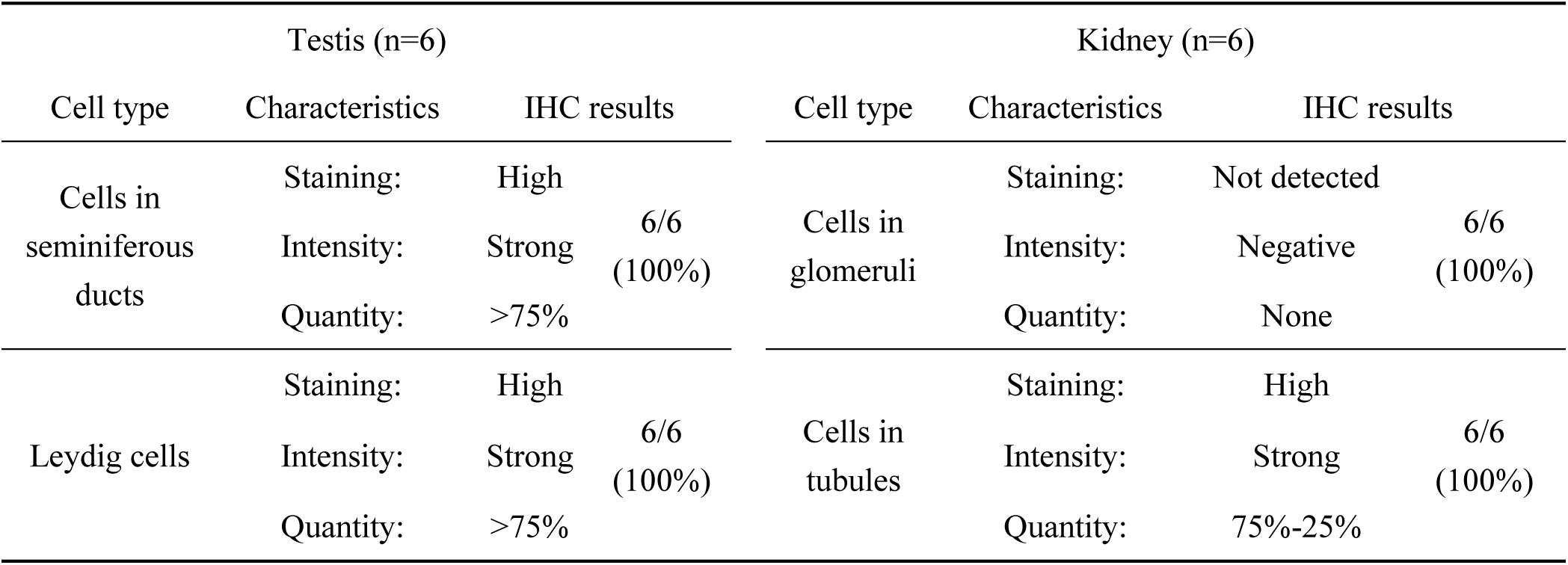
Summary of the IHC results in The Human Protein Atlas project

| Testis (n=6) |  |  |  | Kidney (n=6) |  |  |  |
| --- | --- | --- | --- | --- | --- | --- | --- |
| Cell type | Characteristics | IHC results |  | Cell type | Characteristics | IHC results |  |
| Cells in seminiferous ducts | Staining: | High |  | Cells in glomeruli | Staining: | Not detected |  |
|  | Intensity: | Strong | 6/6 (100%) |  | Intensity: | Negative | 6/6 (100%) |
|  | Quantity: | >75% |  |  | Quantity: | None |  |
| Leydig cells | Staining: | High |  | Cells in tubules | Staining: | High |  |
|  | Intensity: | Strong | 6/6 (100%) |  | Intensity: | Strong | 6/6 (100%) |
|  | Quantity: | >75% |  |  | Quantity: | 75%-25% |  |

**Fig. 2.**
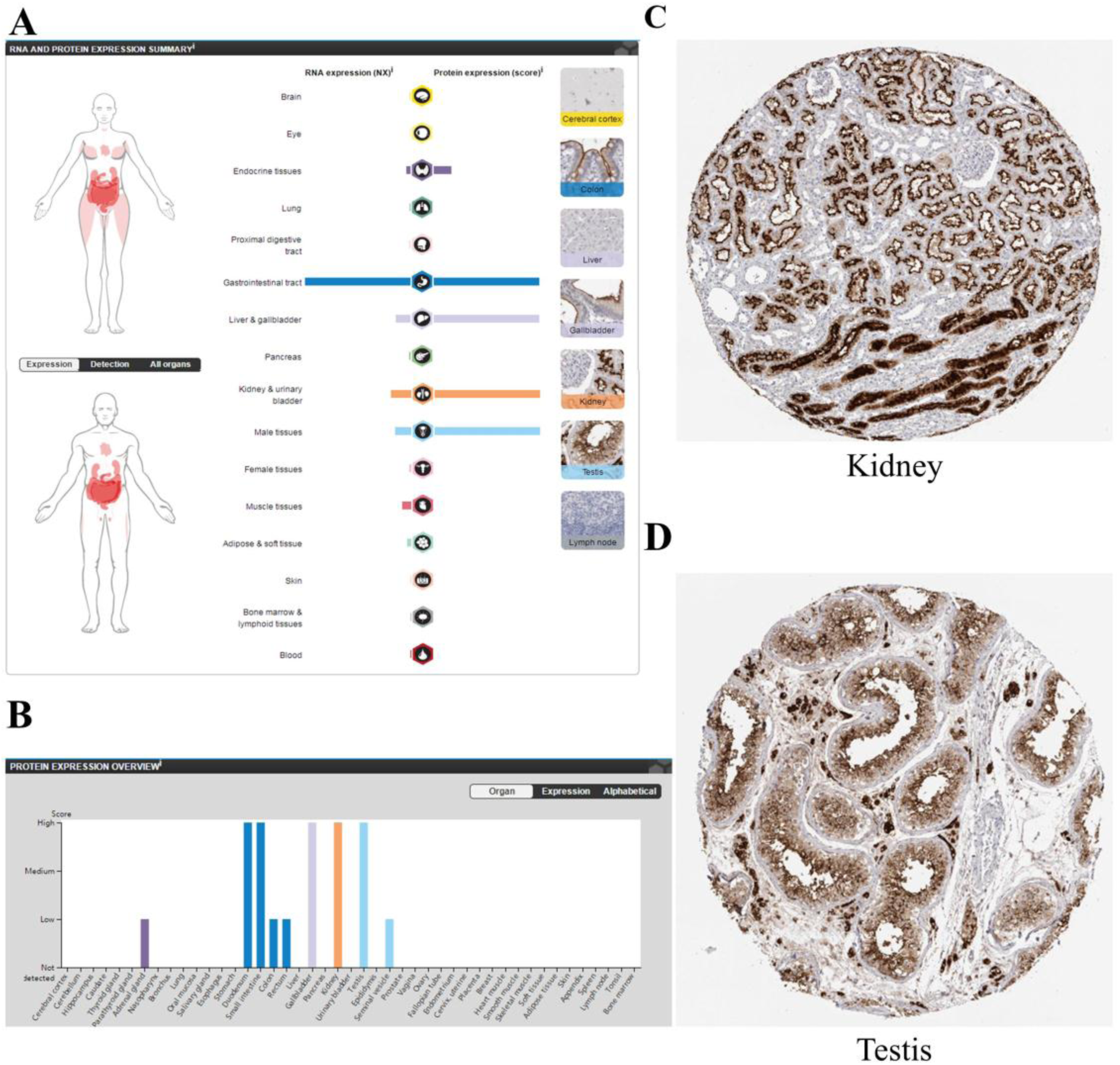
Data of ACE2 protein expression level in different human tissues from HPA portal. A. B. ACE2 protein expression level in different tissues. C. D. Representative IHC staining of ACE2 in kidney and testis.

To further assess the cell type specific expression of ACE2 and confirm ACE2 expression level in kidney, we downloaded the gene expression data of single-cell RNA sequencing of human kidney from Gene Expression Omnibus (GEO) datasets. We first analyzed GSE131685, a published dataset containing scRNA-seq data of the normal kidney samples from 3 donors [17]. We divided single cells into subclusters based on the canonical markers and cell classification in the original literature (Figure 3A), and found specific ACE2 expressions in tubular cells. In contrast, ACE2 expression was not observed in immune cells and glomerular parietal epithelial cells (Figure 3 B). Results of GSE132023 confirm the previous results (data not shown).

Therefore, ACE2 expression in renal tubular cells and testicular cells may suggest a potential mechanism of infection and direct damage of renal tubules and testis by 2019-nCoV binding ACE2 as host cell receptors.

**Fig. 3.**
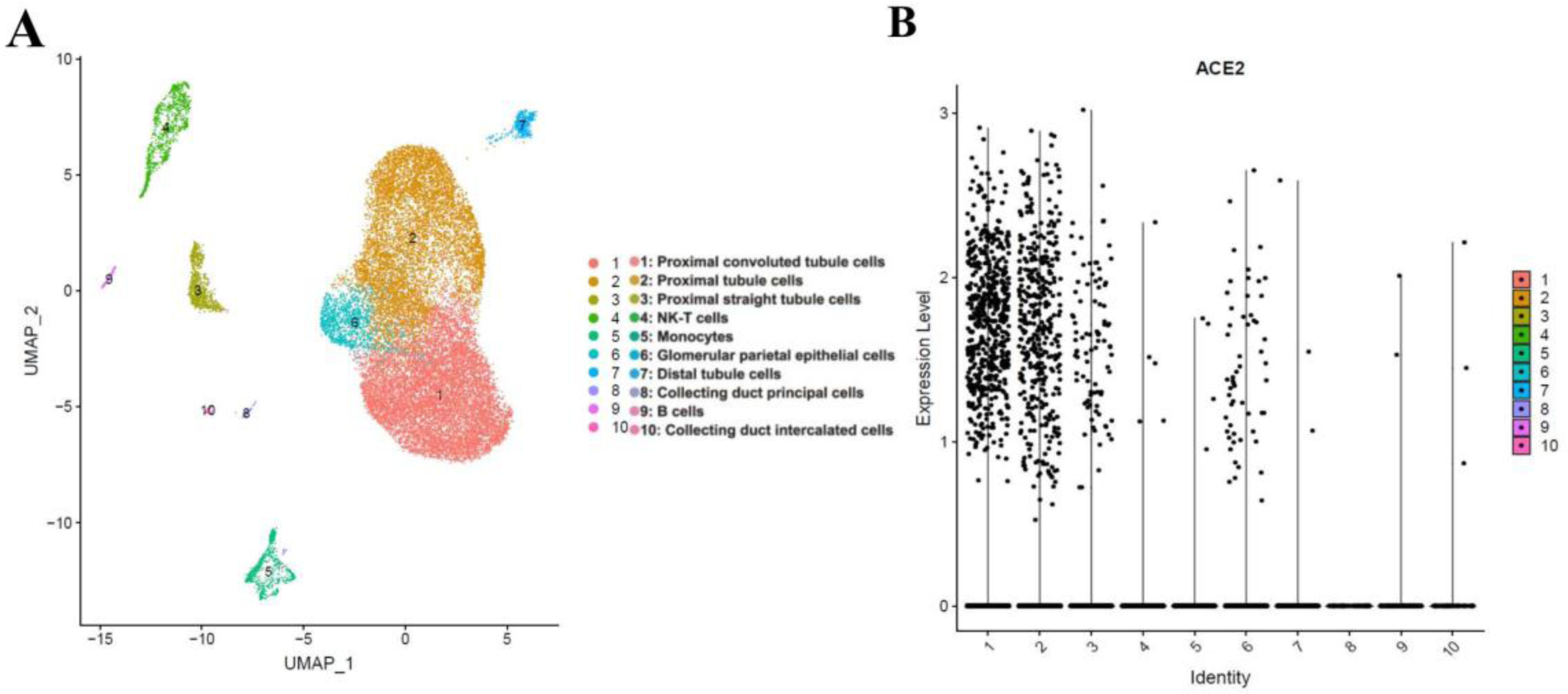
Single cell analysis of published kidney cell atlas. A. Kidney cell atlas visualized by UMAP, colored by cluster number. Cluster number information is provided by the authors. B. The dotplot showing ACE2 gene expression of all major cell types.

## Discussion

Novel coronavirus-infected pneumonia outbroke in Wuhan in December 2019 and January 2020, which has posed a major threat to global public health [18]. The numbers of confirmed cases and deaths are still rising quickly, posing higher challenges for disease control and patient treatment. The symptoms of the disease are complex. In addition to common respiratory symptoms such as cough and fever, some patients may also experience other symptoms such as diarrhea and liver damage [4], or even asymptomatic, which brings more challenges to the patient’s diagnosis and treatment. Studies on the mechanisms of disease pathogenesis could help us understand the disease comprehensively.

Here in this study, we first reviewed the latest literatures and found about 10% of the patients infected with 2019-nCoV had abnormal renal function. This indicates the significance to explore the mechanisms by which the virus affects renal cells. We used the online datasets and bioinformatic methods and found out that ACE2, one of the major receptors for 2019-nCoV, expresses quite highly in renal cells, particularly in tubular cells. IHC results showed no ACE2 expression in cells in glomeruli. Renal tubular cells have reabsorption and excretion functions, and play a key role in excretion of metabolites, maintenance of body fluid balance and acid-base balance. Renal tubular cell injury could cause renal tubules atrophy, thereby aggravating renal interstitial fibrosis, secreting a variety of chemokines and growth factors into the stroma, promoting interstitial inflammatory cell infiltration, interstitial intrinsic cell proliferation, and extracellular matrix (ECM) accumulation. Therefore, 2019-nCoV could enter the renal tubular cell by binding to ACE2, which induces cytotoxicity and abnormal renal function. Examination and follow-up of the renal function of patients infected with 2019-nCoV is necessary to detect the impaired renal function in time and provide early intervention.

Another major point in this study is the high expression level of ACE2 in testicular cells. It is well known that viruses such as HIV, HBV and mumps could enter the testicular cells and cause viral orchitis. Besides, in some cases, virus-induced testicular tissue damage might result in male infertility and testicular tumor [19]. SARS-CoV is just like the ‘cousin’ of 2019-nCoV and shares the receptor ACE2 with 2019-nCoV. Previous research has also investigated the possible damage of the testis in SARS patients and the effects of SARS on spermatogenesis. Their findings suggested that orchitis is a complication of SARS and that spermatogenesis could be affected after infection [20]. Current clinical data show that a large proportion of the novel coronavirus (2019-nCoV) - infected pneumonia patients are young adults and even children, so the potential testicular damage caused by the virus may exist as a late complication. However, limited information is available regarding the involvement of reproductive organs in patients infected with 2019-nCoV. Therefore, our findings suggest that clinicians should take care of the possible occurrence of orchitis. Following-up and evaluation of the reproductive functions should be done in recovered male SARS patients, especially the young male patients.

## Conclusions

Our study demonstrated the highly expression of ACE2 in kidney and testicular tissue and facilitated the understanding of the mechanisms of abnormal renal function and kidney damage in 2019-nCoV-infected patients. Our findings also suggest the patient cares regarding the possible occurrence of orchitis. Following-up and evaluation of the reproductive functions may be necessary in recovered male SARS patients, especially the young male patients.

## Data Availability

In this article, we made use of some online renal single cell RNA-seq (scRNA-seq) gene expression data sets that were publicly usable. These contained the data reported in GSE131685 and GSE107585.
We used RNA and protein expression data of ACE2 in different human tissues and cancer cell lines through The Human Protein Atlas portal (Website: http://www.proteinatlas.org/), GTEx portal (Website: https://gtexportal.org) and The Cancer Cell Line Encyclopedia (CCLE) . All data are available directly online.

https://gtexportal.org

http://www.proteinatlas.org/

